# Emerging SARS-CoV-2 Lineages in Middle Eastern Jordan with Increasing Mutations Near Antibody Recognition Sites

**DOI:** 10.1101/2021.02.09.21251052

**Authors:** Rima Hajjo, Dima A. Sabbah, Sanaa K. Bardaweel

## Abstract

The genomic analysis of the 556 viral sequences from Jordan uncovered three dominant genetic SARS-CoV-2 lineages that are currently circulating in Jordan: B.1.1.312 (76%), B.1.36.10 (11%), and B.1.1.7 (6%), replacing the genetic strains that were dominant before sustained community transmission in Jordan. This raises speculations about these new genetic lineages and their relationship to the severity of disease symptoms in Jordan.

---

Tracking SARS-CoV-2 single nucleotide variants (SNVs) over time and across different geographies is of paramount significance to understand the pandemic dynamics in depth, design better diagnostics and develop effective vaccines. SNVs data is used to construct a phylogenetic tree that to trace changes to the origin (*i*.*e*., reference sequence that first appeared in Wuhan, China). In order to identify the dominant SARS-CoV-2 genetic lineages in Jordan, we downloaded 581 complete viral genomes from the Global Initiative on Sharing All Influenza Data (GISAID)(1) database (http://gisaid.org) on 31 January 2020 and estimated a maximum likelihood tree for these data. We processed the data according to the Methods described by Rambaut *el al*(2), and retained 556 sequences, which had at least 95% coverage of the reference genome (WIV04/MN996528.1) after trimming the 5′- and 3′-untranslated regions, and excluding sequences with > 5% ambiguous base calls (Ns). All Viral lineages were defined by pangolin(3). In order to identify SNVs on the nucleotide and amino acid level, each nucleotide sequence was aligned to the reference genome using CoVsurver(4).

Our analysis revealed that the viral genetic variant landscape is constantly changing with time. New viral clades and lineages appeared in Jordan during the last few months such as the GR strain which now constitutes > 85 % of all analyzed viruses from Jordan. However, some clades which were dominant during the initial weeks of the pandemics such as the V, S and G strains have vanished or became insignificant in numbers. The same observation is true for lineages; lineage B.1.1.312 appeared first in August 2020, and is now the most widely spread viral lineage in Jordan based on public data available at GISAID, and lineage B.1.1.7 is continually growing (Figure 1A). However, lineages such as B.28, B.1.457 and B.1 continue to decline. Our analysis also confirmed the presence of 37 viruses of the B.1.1.7 lineage known as UK variant VUI-202012/01 that appeared in the GR clade and contained the N501Y mutation in the receptor binding domain. Some viruses had concerning combinations of these mutations with deletions altering the spike protein surface.

**Figure 1.**
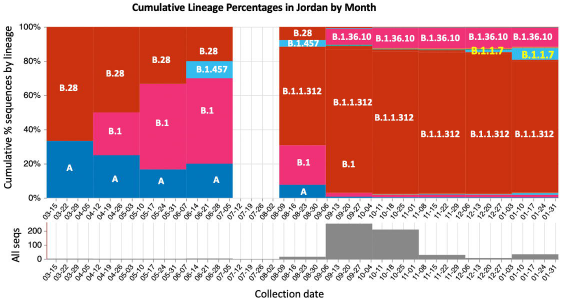
Abundant SARS-CoV-2 lineages and amino acid mutations in spike protein of viruses from Jordan. A) Cumulative lineage percentages in Jordan by month. B) Amino acid mutations in RBD of spike protein(5). All amino acids are colored according to ClustalX properties. C) The 3-dimensional structural model of SARS-CoV-2 spike protein (grey part) in complex with ACE2 (green part) based on PDB 6acj, and with mutations highlighted.

Other amino acid (AA) mutations have been identified in the spike protein of viruses from Jordan over time, however, 10 mutations in particular (Figure 1B) were more concerning than others since they happened in the receptor binding domain (RBD) of the spike protein (Figure 1C), which is fundamental for SARS-CoV-2 entry to human cells (*i*.*e*., increased or decreased ability to enter human cells), antibody recognition and vaccine development. Other mutations that appeared in the RBD of viruses from Jordan, but disappeared from most recent viruses, were N481K and N501I. Mutation N501I also occurred in an important amino acid that is involved in antibody recognition. Although this amino acid is also mutated in the genetic strains of the UK and South Africa, but the Jordanian mutation resulted in a different amino acid substitution from asparagine (N) to isoleucine (I) instead of tyrosine (Y) that is found in 20I/501Y.V1 (B.1.1.7 lineage) of the UK, and 20H/501Y.V2 (B.1.351 lineage) of South Africa. However, evidence is lacking whether the amino acid change from N to I results in the same phenotype caused by Y substitutions. Since both N and Y are polar uncharged amino acids, they may bind to antibodies similarly. However, isoleucine is a non-polar/hydrophobic amino acid that may affect binding interactions with antibodies. These hypotheses should be studied further for their importance.

Interestingly, other mutations that have been recently flagged by GISAID and the scientific community as critical mutations such as: S477N (part of large Melbourne outbreak from clade GR and some Central European clade GH clusters), N439K (long lasting UK outbreak with clade G and European spillover), and Y453F (mink adaptation and part of limited outbreaks) were not identified in the viruses from Jordan. All details on materials, methods, genomic analysis and mutational results are provided in Supporting Information and in Supplemental Tables S1 and S2. All raw data and results files are deposited on GitHub and available for researchers to access (https://github.com/rhajjo/Hajjo_EID.git).

According to daily official reports published by the Jordanian Ministry of Health, Jordanians were showing more disease symptoms starting from September 2020 with increased need to intensive care and/or ventilators. Jordan also witnessed an increased disease transmission during September through December of 2020. This was accompanied by changes in the clade and lineage during that time too; suggesting that emergent clades and lineages could have changed the virulence and pathogenicity of the virus, or causing different clinical presentation. Nonetheless, this remains an interesting observation that requires follow up studies to verify whether the increased disease transmissibility during the Fall and early Winter has been a mere result of both societal behaviors and governmental relaxed control measures, or due to the appearance of specific viral clades and lineages.

## Data Availability

Raw data files and results of the genomic analyses are provided on github. The authors will make these data publicly available and supply additional details and a readme files upon request.

https://github.com/rhajjo/Hajjo_EID.git

## Acknowledgments

We gratefully acknowledge all the Authors from the Originating laboratories responsible for obtaining the specimens and the Submitting laboratories where genetic sequence data were generated and shared via the GISAID (http://www.gisaid.org) Initiative, on which this research is based. RH and DS acknowledge support from the Deanship of Scientific Research at Al-Zaytoonah University of Jordan (Grant number 2020-2019/17/03).

Dr. Hajjo is an Assistant Professor of Pharmacogenetics and Pharmacoinformatics at Al-Zaytoonah University of Jordan, an Adjunct Professor at UNC-Chapel Hill, and a Board Member at the National Center for Epidemics and Communicable Disease Control in Amman, Jordan.

## Author Contributions

RH generated the idea and performed formal analysis of epidemiology and genetic data, and wrote the manuscript. DS and SB participated in data procurement, in critical scientific discussions, and helped in manuscript edits.

## Data Availability

Raw data files and results of the genomic analyses are provided on github. The authors will make these data publicly available and supply additional details and a readme files upon request. https://github.com/rhajjo/Hajjo_EID.git

## Notes

The authors declare no competing financial interest.

## Supporting Information

Supplementary Information for materials and methods.

Supplementary Table S1. Genomic analysis results from CoV-Glue.

Supplementary Table S2. The complete list PANGOLIN lineages assigned to query sequences.

---

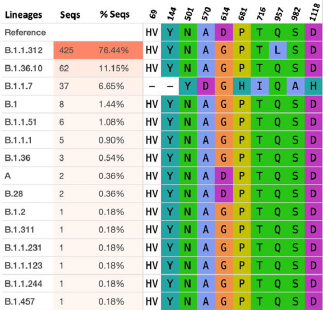

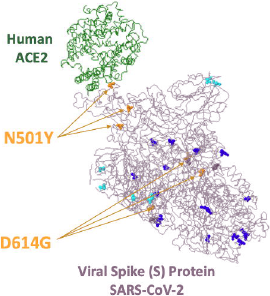

