## Supporting Information for "Emerging SARS-CoV-2 Lineages in Middle Eastern Jordan with Increasing Mutations Near Antibody Recognition Sites"

**Materials and Methods**

**Epidemiology and genomic data.** COVID-19 epidemiology data have been retrieved from the WHO COVID-19 dashboard(1) and the Jordanian Ministry of health website(2). All 556 genomic sequences for SARS-CoV-2 viruses found in Jordan were downloaded from GISAID(3).

**Genomic analysis.** CoV-GLUE was used for the analysis and interpretation of SARS-CoV-2 virus genomic sequences, with a focus on amino acid sequence variation. This freely available web application is based on the GLUE(5) environment and provides a browsable database of amino acid replacements and coding region indels that have been observed in sequences from the COVID-19 pandemic. Pair-wise alignments were also performed for all was performed to all SARS-CoV-2 virus sequences of interest using MAFFT(6). After excluding irrelevant sequences and those with potential quality issues, amino acid replacements and in-frame indels in each sequence were identified.

**Mutational analysis.** CoVsurver(6) from GISAID was used in order to identify to identify candidates for phenotypic changes of special epidemiological relevance. Additionally, this application enabled the visualization of the position of the relevant mutation(s) in structural models and highlighted whether these mutations are close to common host receptor or antibody binding sites.

**Phylogenetic assignment of named global outbreak lineages.** Phylogenetic Assignment of Named Global Outbreak LINeages (PANGOLIN)(7) version v3.0 was used to assign lineages to query sequences downloaded from GISAID. This software was developed by the Centre for Genomic Pathogen Surveillance. All methods implemented in this software are described in Rambaut *et al* 2020(8).

**Clade and lineage nomenclature.** We used GISAID’s nomenclature system for classifying major clades. Clade assignments are based on genetic markers (i.e., marker mutations) within seven high-level phylogenetic groupings from the early split of S and L clades, to the further evolution of L into V and G and later of G into GH, GR and GV (cf. Table 1). Additionally, GISAID clades were also described in details based on lineages assigned by PANGOLIN, which allows for a better understanding of the global spread of COVID-19.

**Data visualization.** Jalview(9) v.2.11.1.3, CoVsurver and COVID CoV Genomics(10) tools were used to analyze viral sequences and to identify mutations from the multiple sequence alignment results.
